# Incidence and outcome of delirium during Helmet CPAP treatment in COVID-19 patients

**DOI:** 10.1101/2021.05.01.21256071

**Authors:** Federica Samartin, Emanuele Salvi, Anna Maria Brambilla, Alessandro Torre, Stella Ingrassia, Antonio Gidaro

## Abstract

It is estimated that almost one-third of patients with COVID-19 develop delirium in the course of disease, actually it may be the only presenting symptom, especially in dementia patients. In COVID-19 patients delirium is associated with higher mortality rate, increased length of stay and a greater rate of admission in Intensive Care Unit and ventilator utilisation. We hypothesized a greater rate of delirium in Helmet CPAP COVID-19 ventilated patients because many known risk factors for delirium co-exist in these kind of patients (i.e. isolation, noise, dehydration). The first aim of our study is to investigate the incidence of delirium occurring during Helmet CPAP therapy in COVID-19 patients. Moreover, we wanted to verify if there are predictable risk factors for delirium and to determine if delirium increases the risk of adverse outcomes (need of endotracheal intubation and death). The cohort of CPAP ventilated COVID-19 patients were composed by 194 patients. Of them, 57 patients (29.3%) developed delirium during CPAP, more than two third in the first 48h. Age over 70 years, previous diagnosis of dementia or psychiatric condition, P/F < 150 after starting CPAP and Gr/Lys >8 resulted risk factors for delirium. Delirium group had a significantly higher mortality rate (47% vs 23%) and lower intubation rate (12% vs 26%) compared to non-delirious ones. Despite many potential predisposing factors are common in CPAP ventilated patients, delirium incidence in our population seems not to differ from what reported by other studies. Moreover, the occurrence of delirium seems not to be related to prolonged CPAP treatment, indeed no correlation between time spent in CPAP and delirium onset was found.

## Manuscript

Coronavirus disease 2019 (COVID-19) is an infectious disease caused by severe acute respiratory syndrome coronavirus 2 (SARS-CoV-2) which may causes Acute Hypoxemic Respiratory Failure (AHRF) or Acute Respiratory Distress Syndrome (ARDS) requiring Non-Invasive Ventilation (NIV) and in particular Helmet Continuous Positive Airway Pressure (CPAP), preferred for use both as a ventilator and as a protection against viral transmission for healthcare workers.

Beyond the classical signs and symptoms of presentation as fever and cough, it is estimated that more than one-third of patients with COVID-19 develop neurologic or neuropsychiatric symptoms that seem to be associated with more severe disease [1]. Although neurologic manifestations can happen in a broad spectrum of acute and infectious diseases, there are evidences suggesting that the coronavirus family is specially neurotropic, indeed during Severe Acute Respiratory Syndrome (SARS) and Middle Est Respiratory Syndrome (MERS) epidemics, many neurological complications were described and signs suggestive of delirium are common in the acute stage of SARS, MERS and even COVID-19 [2]. Delirium should be recognized as a potential feature of COVID-19 and may be the only presenting symptom, especially in dementia patients [3].

Occurring of delirium has a prognostic significance, in COVID-19 patients delirium has been reported to be independently associated with higher mortality rate, increased length of stay and a greater rate of admission in Intensive Care Unit (ICU) and ventilator utilisation [3-5].

Although no data are available about a direct correlation between NIV and delirium, several well-known precipitating factors of delirium can co-exist during NIV, especially with helmet interface, as. So much that it might be possible that prolonged CPAP could increase delirium rate.

The first aim of our study is to investigate the incidence of delirium occurring during Helmet CPAP therapy in COVID-19 patients. Moreover, we wanted to verify if there are predictable risk factors for delirium in CPAP ventilated patients and to determine if delirium increases the risk of adverse outcomes, defined as need of endotracheal intubation (ETI) and death. Secondly, we better characterize delirium’s risk factors including them in a Cox proportional hazard multivariate model.

We analysed data from patients admitted to the internal medicine and infectious diseases wards of “Luigi Sacco” University Hospital of Milan from 21 February to 5 May 2020.

The study is part of a prospectively conducted registry study (“REGISTRO DELLE INFEZIONI SOSPETTE E ACCERTATE COVID-19/Studio Sacco COVID-19)” was approved by the local ethical committee Milan Area 1 in ASST Fatebenefratelli Sacco, University HospitalLuigi Sacco with the registration number 2020/16088.

### The inclusion criteria were

1. COVID-19 pneumoniae defined by positivity of real-time reverse transcription-polymerase chain reaction test (RT-PCR) at nose-pharyngeal swab for SARS-CoV-2 and a Chest X-Ray positive for interstitial pneumonia.
2. AHRF requiring Helmet CPAP treatment (PaO2/FiO2 ≤ 200 mmHg and respiratory rate >25/minute).

### DSM-5 criteria were used for the diagnosis of delirium

1. presence of disorder of consciousness with reduced ability to focus, sustain, or shift attention.
2. change in cognition that is not better accounted for by a pre-existing, established, or evolving dementia.
3. development of the disorder over a short period with fluctuation during the day.
4. evidence that the disorder is caused by a direct physiologic consequence of a medical condition.

Kolmogorov-Smirnov test was used to evaluate the normality of data distribution. Qualitative data were expressed as number and percentage. Chi square or Fisher exact tests were used for group’s comparison. Quantitative data were expressed as mean, standard deviation, median and range. Student T-test and Mann-Whitney test (for non-parametric data) were used for comparison between groups. *P*-value less than 0.05 was considered statistically significant. Cox proportional hazard model was used to evaluate four delirium risk factors: age, previous diagnosis of dementia or psychiatric condition, P/F soon after starting CPAP and Gr/Ly. ***Excel*** (Office program 2010) and ***SPSS*** (statistical package for social science-SPSS, Inc., Chicago, IL version 26) were used for statistical analysis.

During the observational period 1016 COVID-19 patients were admitted to our Hospital; 194 (19,1%) met the inclusion criteria. Their clinical characteristics are resumed in Table 1. Of them, 57 patients (29.3%) developed signs and symptoms of delirium during CPAP. 32 patients (56.1%) developed delirium during the first 24 hours after starting CPAP and 48 (84.2%) in the first 48 hours.

**Table 1.** Clinical and biochemical characteristics of examined population. T1 = first data after starting CPAP. CCI= Charlson Comorbidity Index

|  | *GENERAL*<br>*POPULATION* | *NO*<br>*DELIRIUM*<br>*137*<br>*70.6%* | *DELIRIUM*<br>*57*<br>*29.4%* | *P VALUE*<br>*UNIVARIATE* | *ODDS RATIO*<br>*UNIVARIATE* |
| --- | --- | --- | --- | --- | --- |
| *Age (years)* | 64.0 ± 14.7 | 60.9 ± 14.4 | 71.5 ± 12.7 | **<0.001** |  |
| *Age >70years* | 77 (36,7%) | 40 (29,2%) | 37 (64,9%) | **< 0.0001** | **4,5 (2,3 to 8,7)** |
| *Male Sex* | 139 (71.6%) | 100 (73.0%) | 39 (68.4%) | 0.52 |  |
| *CCI* | 2.66 ± 2.23 | 2.21 ± 1.98 | 3.76 ± 2.45 | 0.121 |  |
| *Dementia or previous*<br>*psichiatric condition* | 19 (9.8%) | 9 (6.6%) | 10 (17.5%) | **0.024** | **3 (1.2 to 7.9)** |
| *PaO2/FiO2 at T1* | 163 ± 84 | 173 ± 84 | 140 ± 81 | **<0.001** |  |
| *PaO2/FiO2 at T1*<br>*<150* | 99 (51%) | 60 (43,7%) | 39 (68,4%) | **0,002** | **2,8(1,5 to 5,3)** |
| *Time in CPAP (hours)* | 153.8 ± 134.4 | 147.7 ± 115.8 | 168.4 ± 171.4 | 0.336 |  |
| *GRs/Ly ratio* | 9.1 ± 9.0 | 7.98 ± 8.21 | 11.64 ± 10.23 | **0.011** |  |
| *GRs/Ly ratio > 8* | 55 (28,4%) | 24 (17,5%) | 31 (54,9%) | **0.004** | **2,6 (1,4 to 4,9)** |
| *CRP (mg/L)* | 165 ± 95 | 157 ± 90 | 183 ± 104 | 0.088 |  |
| *D-dimer* | 8422 ± 21728 | 6493 ± 19545 | 13748 ± 26396 | 0.109 |  |
| *IL-6* | 139 ± 377 | 132 ± 379 | 161 ± 377 | 0.698 |  |
| *Steroids therapy* | 19 (9.8%) | 10 (7.3%) | 9 (15.8%) | 0.07 |  |
| *Lenght of stay (days)* | 20.6 ± 14.6 | 20.8 ± 15.0 | 19.1 ± 13.0 | 0.52 |  |
| *Deaths* | 59 (3.4%) | 32 (23.4%) | 27 (47.4%) | **<0.001** | **3 (1,5 to 5,7)** |
| *ETI* | 42 (21.6%) | 35 (25.5%) | 7 (12.3%) | **0.041** |  |

Patients developing delirium were older, had a higher frequency of previous diagnosis of dementia or psychiatric condition, a more compromised respiratory function as assessed by PaO2/FiO2 ratio (immediately after starting CPAP) and a higher GRs/Ly ratio. No significant difference between the two groups were found with respect to comorbidity burden (CCI), biochemical inflammation-related parameters (IL-6, CRP, D-dimer) nor the use of corticosteroids. The length of hospitalization did not differ, being approximately 20 days in both groups. Time spent in CPAP was highly variable and no statistical significance has been found between the two groups.

Twenty-seven out of 57 patients (47%) presenting delirium died, accounting for a higher mortality rate in this group. The intubation rate was lower in the delirium group than in patients that didn’t develop delirium. Noticeably, delirious patients who underwent ETI were younger than non-intubated delirious ones (61.4±13.4 vs 72.9±12, p 0.02) and with a trend to a lower comorbidity burden [Charlson Comorbidity Index <4 in 5/7 (83%) of intubated patients vs 19/50 (43%) in non-intubated patients, p 0.06].

At Cox proportional hazard model, resumed in Table 2, age (p 0.04) and PaO2/FiO2 ratio after starting CPAP (p 0.02) were significantly correlated with delirium presentation, while GRs/Ly ratio and previous diagnosis of dementia or psychiatric conditions didn’t reach statistical significance.

**Table 2.** Cox proportional hazard model: age, previous diagnosis of dementia or psychiatric condition, P/F soon after starting CPAP and Gr/Ly

|  | *P VALUE MULTIVARIATE* | *ODDS RATIO MULTIVARIATE* |
| --- | --- | --- |
| *Age (years)* | **0.04** | **1.025** |
| *Dementia or previous*<br>*psichiatric condition* | 0.145 | 1.718 |
| *PaO2/FiO2 at T1* | **0.02** | **0.996** |
| *GRs/Ly ratio* | 0.06 | 1.024 |

The main finding of our work is that almost one third (29.3%) of a population of consecutive Helmet CPAP ventilated COVID-19 patients presented delirium; this complication was associated with a significantly higher mortality, according to the literature [3-5].

Despite many potential predisposing factors are common in CPAP ventilated patients with AHRF-as hypoxia, dehydration, isolation and noise-delirium incidence in our population seems not to differ from what reported by other studies conducted in populations in which ventilated patients were not represented [3] or constituted a minority of the studied subjects [4,5]. Moreover, the occurrence of delirium seems not to be related to prolonged CPAP treatment, as in more than 80% of the patients it developed in the first 48 hours and no correlation between time spent in CPAP and delirium onset was found.

Our data confirm that age and previous diagnosis of dementia or psychiatric condition can be considered risk factors for the development of delirium [3,4]. Furthermore, severe respiratory impairment is associated to delirium in our cohort as PaO2/FiO2 ratio was statistically lower in delirum patients. This latter finding is in accordance with previous work reporting an increased risk for neuropsychiatric symptoms in patients with more severe COVID-19 disease and worst respiratory failure [1].

We observed a significantly lower intubation rate in patients who developed delirium; in fact delirious patients had higher mean age and the higher prevalence of dementia, possibly limiting the indication to intubation.

The study presents some limitations. First, it is possible that patients at greater risk for delirium (i.e. elderly with severe dementia) were not ventilated, thus making possible an under-estimation of delirium occurrence when compared to non-ventilated patients. Moreover, populations in comparison found in literature were not completely comparable to ours for age, severity of illness and settings. For these reasons this data should be taken carefully. For last, our study was performed in a single center dedicated to high-complexity medical care and our results should not be generalized to different populations.

In conclusion CPAP therapy seems not to increase the delirium occurrence, as indicated by an incidence in our cohort of non-invasive ventilated patients similar to what found in studies regarding non-ventilated COVID-19 subjects. Unfortunately, COVID-19 pandemic, especially during the first wave, drastically challenged hospital organization, forcing to the use of Helmet CPAP also outside ICU or sub-intensive units, where monitoring patients is more difficult. Our study may add evidences for the recognition of patients at high risk for delirium (over 70 years; mentally ill; GRs/Ly ratio > 8; P/F < 150 mmHg after starting CPAP) needing a strict supervision and the highest intensity of care setting for at least the first 48 hours.

## Data Availability

All data referred to in the manuscript are available for request

